# Alternative or Complementary Role of Serological Rapid Antibody Test in the Management of Possible COVID-19 Cases

**DOI:** 10.1101/2020.09.13.20193615

**Authors:** Fatma Yildirim, Pınar Yildiz Gulhan, Ozlem Ercen Diken, Aylin Capraz, Meltem Simsek, Berna Botan Yildirim, Muhammet Ridvan Taysi, Sakine Yilmaz Ozturk, Nurcan Demirtas, Julide Ergil, Adem Dirican, Tugce Uzar, Irem Karaman, Sevket Ozkaya

## Abstract

**Background:** Although the gold diagnostic method for COVID-19 is accepted as the detection of viral particles by reverse transcription polymerase chain reaction (RT-PCR), serology testing for SARS-CoV-2 is at increased demand. A primary aim for utilization of serological tests are to better quantify the number of COVID-19 cases including those RT-PCR samples were negative but showing clinical and radiological signs of COVID-19. In this study, we aimed to report the features of the patients that were diagnosed and treated as possible COVID-19 cases whose multiple nasopharyngeal swab samples were negative by RTPCR but serological IgM/IgG antibody against SARS-CoV-2 were detected by rapid antibody test.

**Method:** We retrospectively analyzed eighty suspected COVID-19 cases that have at least two negative consecutive COVID-19 PCR test and were subjected to serological rapid antibody test.

**Result:** The specific antibodies against SARS-CoV-2 were detected as positive in twenty-two patients. The mean age of patient group was 63.2 ± 13.1 years old with male /female ratio 11/11. Cough was the most common symptom with 90.9%. Most common presenting chest CT findings were bilateral ground glass opacities (77.2%) and alveolar consolidations (50.09%). The mean duration from symptom initiation to hospital admission, to hospitalization, to treatment initiation and to detection of antibody positivity were 8.6 ± 7.2, 11.2 ± 5.4, 7.9 ± 3.2 and 24 ± 17 days, respectively.

**Conclusion:** Our study demonstrated the feasibility of COVID-19 diagnosis based on rapid antibody test in the cases of patients whose RT-PCR samples were negative. We suggest that the detection of antibodies against SARS-CoV-2 with rapid antibody test should be included in the diagnostic algorithm in suspected COVID-19 patients.

## Introduction

Novel coronavirus disease (COVID-19) is a unique pneumonia caused by SARS-CoV-2 that typically causes various degrees of respiratory disease.^1^ Currently, the entire world is battling with COVID-19 pneumonia which can be dreadfully lethal in high-risk patient groups. Although COVID-19 diagnosis is generally made based on clinical, laboratory and radiological features of the patients, the most common diagnostic method is the real time reverse transcription polymerase chain reaction (RT-PCR)^2,3^. However, several studies are published about the concerns regarding the sensitivity of RT-PCR tests^4,5^. False negative results are thought to be due to PCR tests itself or errors during performing the test, whereas false positive results are rarely seen.^4^

Tests that produce results in 15 minutes, can detect IgM and IgG antibodies produced against SARSCoV-2, and they have been approved in Europe, as well as in China. Although the accuracy rate of these tests is lower than PCR, it has not been investigated when these tests should be studied or in which cases they would be useful in the diagnosis. In this study; we aimed to investigate whether these rapid antibody tests would be useful in the diagnostic challenge of patients whose PCR tests were negative but has radiologically and clinically consistent features with COVID-19 pneumonia.

## Material and Method

We retrospectively analyzed the patients with COVID-19 who have at least two consecutive PCR test results were negative for COVID-19 and specific antibodies against SARS-CoV-2 were positive. To authors’ knowledge, this is the first case series investigating the diagnostic value of antibody tests in the management of patients with possible COVID-19 pneumonia whose RT-PCR tests were negative.

### Patient Selection

In Turkey, rapid antibody test kits for COVID-19 were commercially available from the beginning of April 2020. To date, we were able to test eighty patients that were suspected as possible COVID-19 cases and detected twenty-two serologically positive cases in our centres. Herein, we introduced features of twenty-two positive cases as a case-series study. All values were given as mean and percent of cases. Symptomatic PCR-negative patients who were suspected to be infected with SARS-CoV-2 based on positive radiological findings were included in the study. High resolution computed tomography (HRCT) was used for the radiological assessment. In patients with COVID-19 pneumonia, ground-glass formation and/or consolidative opacities distributed usually bilateral, peripheral, and mostly basal, were considered as positive HRCT findings. The patients with negative PCR tests were tested for specific antibodies against SARS-CoV-2 following the COVID-19 treatment.

### COVID-19 testing

Samples were taken from the patients with nasopharyngeal and nasal swabs and they were analysed by RT-PCR test. The humoral responses against SARS-CoV-2 were tested with rapid card test with blood samples of patients. The blood taken from the patient was dropped on this rapid card test and the antibody response was checked. The clinical samples that had been deposited in were anonymized and used in accordance with local ethical guidelines. Total antibody levels against SARS-CoV-2 were noted. We used the Colloidal Gold SARS-CoV-2 IgG/IgM Rapid Test (Beijing Hotgen Biotech Co., Ltd) to detect antibodies against the SARS-CoV-2 virus. This test is a lateral flow chromatographic immunoassay. The test cassette consists of: 1) a burgundy colored conjugate pad containing SARS-CoV-2 recombinant antigens (S and N proteins) conjugated with colloidal gold (SARS-CoV-2 conjugates) and rabbit IgG-gold conjugates; 2) a nitrocellulose membrane strip containing an IgG line (G Line) coated with anti-human IgG, an IgM line (M Line) coated with anti-human IgM, and the control line (C Line) coated with goat anti-rabbit IgG. When a correct volume of test specimen is dispensed into the test cassette, the specimen migrates by capillary action along the cassette. The anti-SARS-CoV-2 virus IgG, if present in the specimen, will bind to the SARS-CoV-2 conjugates. If IgG is present in the specimen, the immunocomplex will then captured by the anti-human IgG line, forming a burgundy colored G Line, indicating a SARS-CoV-2 virus IgG positive test result. The anti-SARS-CoV-2 virus IgM, if present in the specimen, will bind to the SARS-CoV-2 conjugates. The immunocomplex is then captured by the anti-human IgM line, forming a burgundy colored M Line, indicating a SARS-CoV-2 virus IgM positive test result.

## Results

Total of twenty-two patients whose nasopharyngeal swab PCR test were negative and clinical findings were positive including chest CT, and symptoms that were supporting the COVID-19 included in the study and the characteristics of patients were shown in Table I. All of the patients’ at least two consecutive PCR tests minimum two days apart were negative. The mean age was 63.2 years old and male to female ratio was 11/11. The mean duration of times from symptoms to hospital admission, hospital stay, drug using and antibody positivity from first symptoms started were 8.6 ± 7.2, 11.2 ± 5.4, 7.9 ± 3.2 and 24 ± 17 days, respectively. The radiological findings and drug regimens were shown in Table II. The radiological findings such as bilateral reticular and ground-glass opacities were demonstrated in Figure 1–5. Also, dense consolidations were noted in Figure 3 and 5. The bilateral fibroreticular infiltrates with crazy-paving patterns were given in Figure 6. The laboratory results of the patients were given in Table III.

**Table I.** Characteristics of Patients

|  | n(%) |
| --- | --- |
| **Age(year)** |  |
| *Mean ±SD* | 63.2±13.1 |
| **Gender** |  |
| *Male/Female* | 11/11 |
| **Symptoms (n, %)** |  |
| *Cough* | 20(90.9) |
| *Dyspnea* | 14(63.6) |
| *Fever* | 10(45.4) |
| *Chest pain* | 8(36.3) |
| **Duration (day/mean ±SD)** |  |
| *from symptom to admission* | 8.6 ± 7.2 |
| *hospital stay* | 11.2 ± 5.4 |
| *from symptoms to antibody test* | 24 ± 17 |
| *drug treatment* | 7.9 ± 3.2 |

**Table II.** The Radiological Findings and Drug Regimens

|  | n (%) |
| --- | --- |
| ***Radiology*** |  |
| *GGO\** | 17(77.2) |
| *Consolidation* | 11(50) |
| *Nodular infiltrates* | 4(18.1) |
| *Fibroreticular infiltrates* | 3(5.9) |
| ***Drug Regimens*** |  |
| *HCQ\*\*+Azithromycine+Favipravir* | 11(50) |
| *HCQ\*\*+Azithromycine* | 7(31.8) |
| *Favipravir* | 4(18.1) |

*\*GGO: Ground-Glass Opacities, \*\*HCQ: Hydroxychloroquine*

**Table III.** The laboratory parameters of patients

| Value | n |
| --- | --- |
| *ESR\** (mm/h) | 68.5 ± 41.7 |
| *LDH\*\** (U/L) | 362 ± 152 |
| *CRP\*\*\** (mg/L) | 95 ± 101 |
| *Ferritine* (µg/l) | 778± 684 |
| *WBCs\*\*\*\** | 8621 ± 3549 |
| *Lymphocytes* (n) | 1430 ± 530 |
| *Lymphocytes* (%) | 22 ± 10.8 |
| *Neutrophils* (n) | 5390 ± 2450 |
| *Neutrophils* (%) | 70.5 ± 12.3 |
| *D-Dimer* (mg/L) | 1875 ± 2757 |
| *Procalcitonin* (mg/L) | 0.15 ± 0.03 |

\**ESR*: Erythrocyte Sedimentation Rate, \*\**LDH*: Lactate Dehydrogenase
\*\*\**CRP*: C-reactive protein, \*\*\*\**WBCs*: White Blood Cells

**Figure 1.**
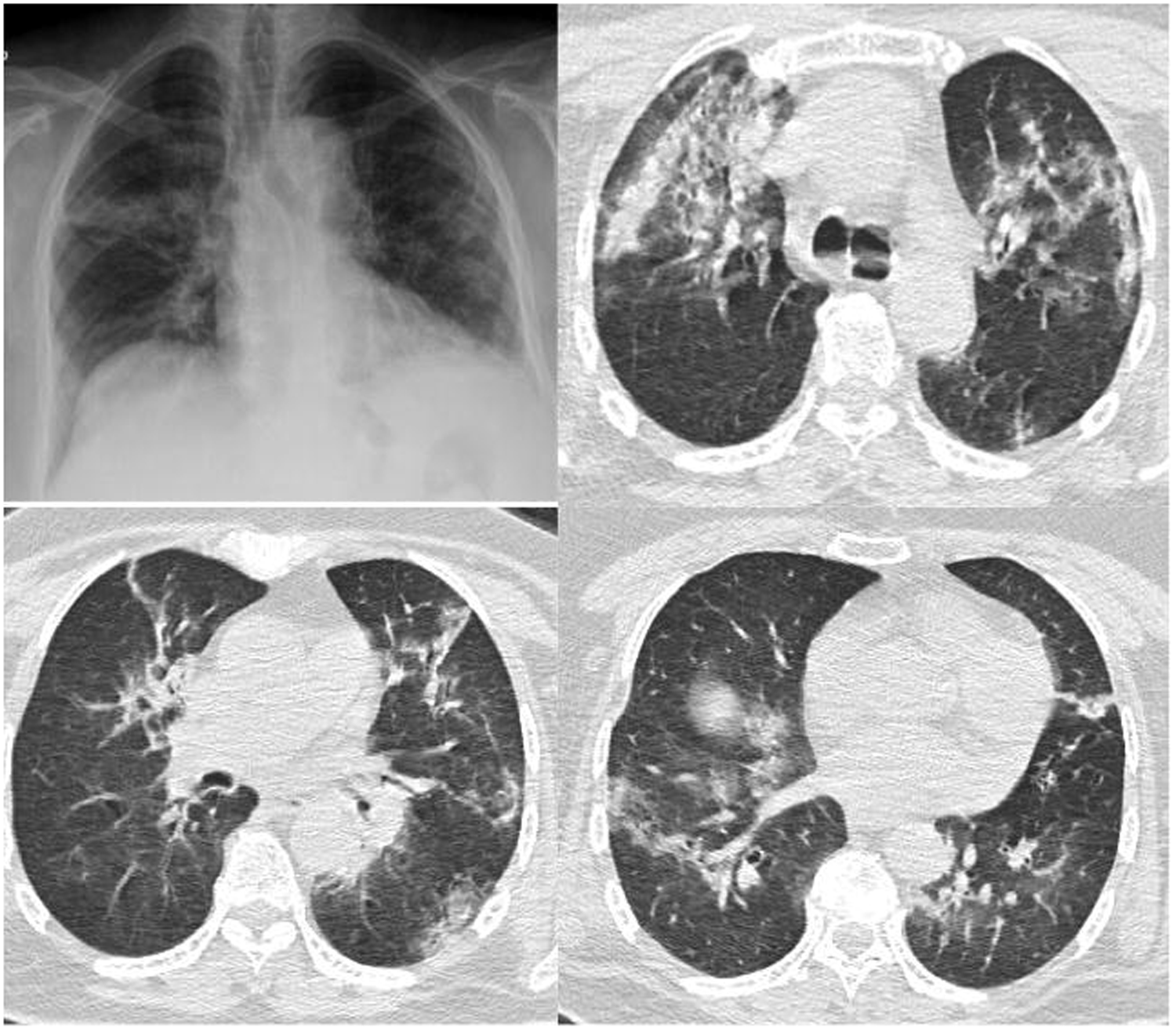
Chest radiograph and HRCT images showing the bilateral reticular and ground-glass opacities.

**Figure 2.**
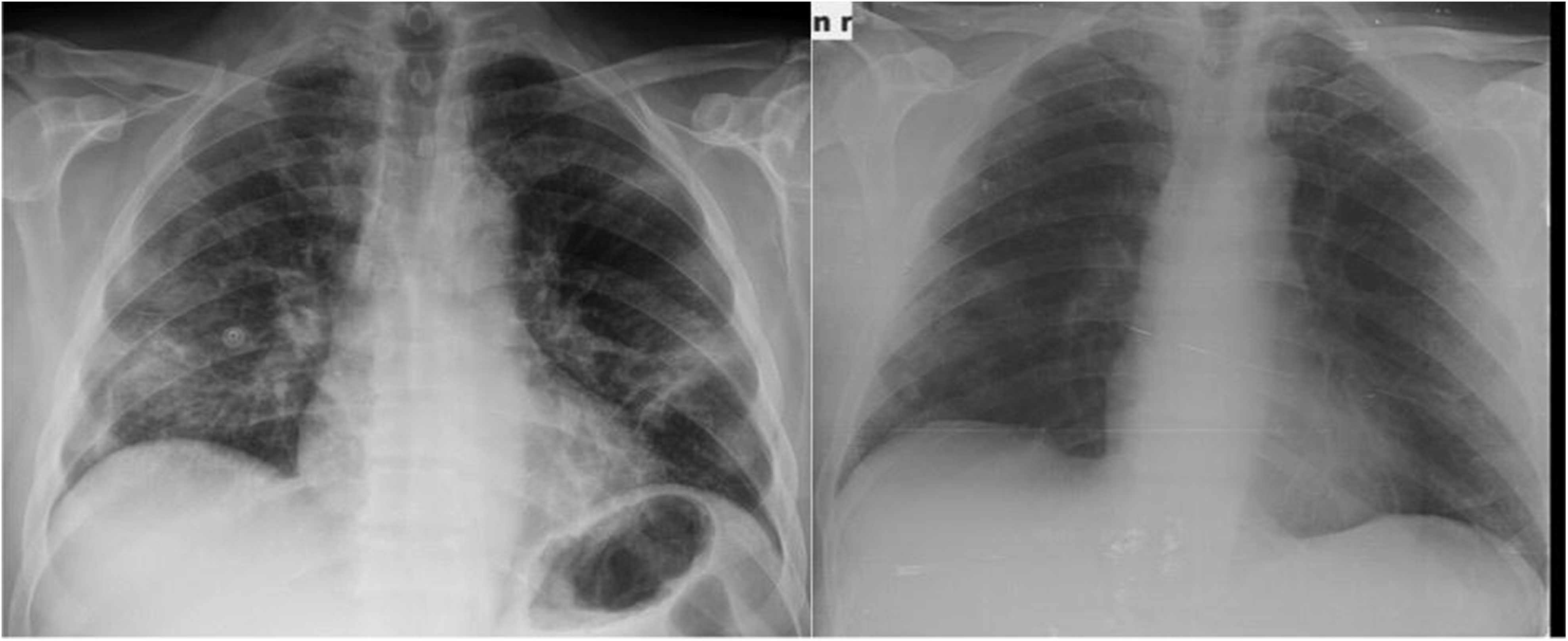
Chest radiographies showing the bilateral infiltrates before (left) and after the treatment (right).

**Figure 3.**
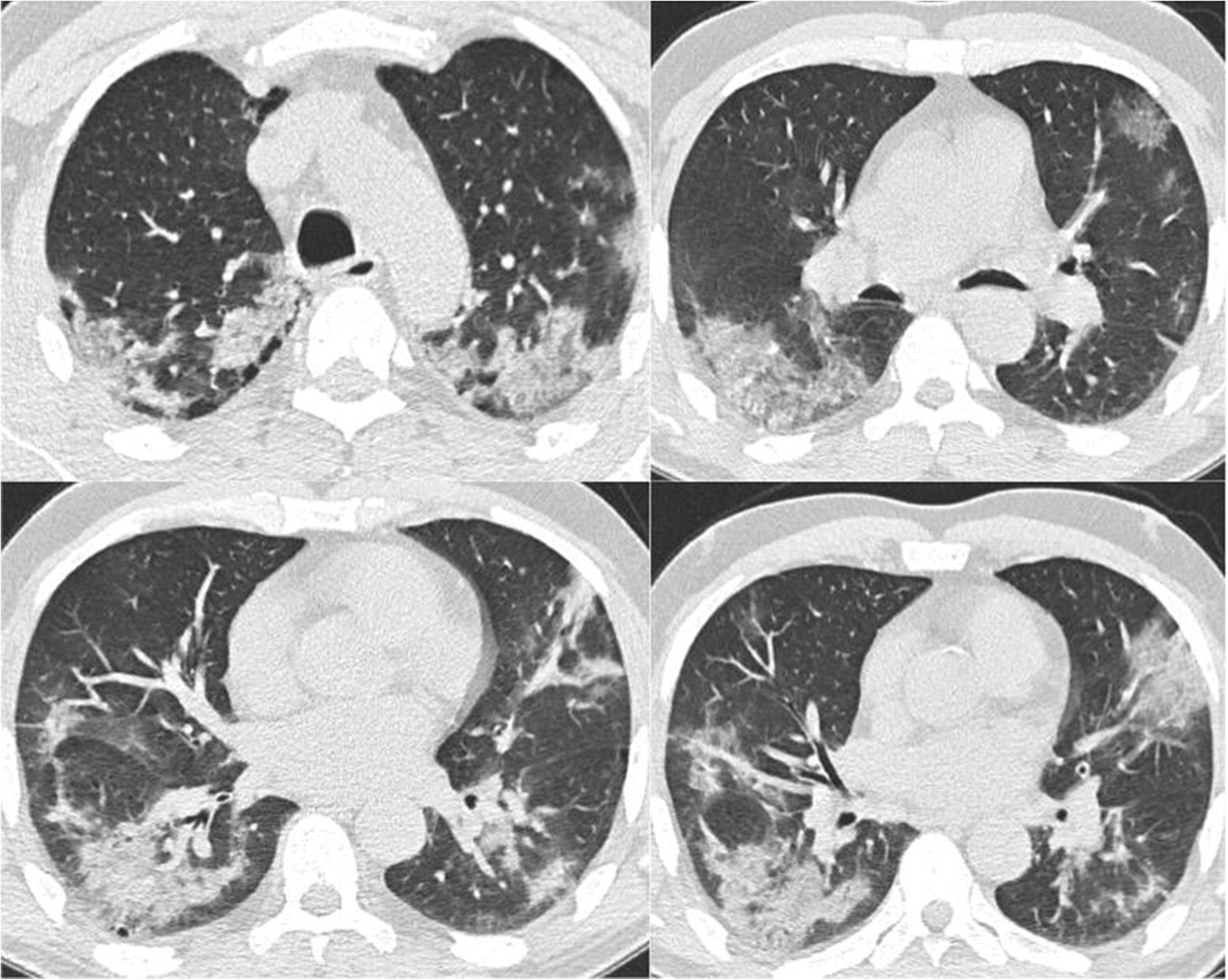
HRCT images showing the bilateral ground-glass opacities and consolidations.

**Figure 4.**
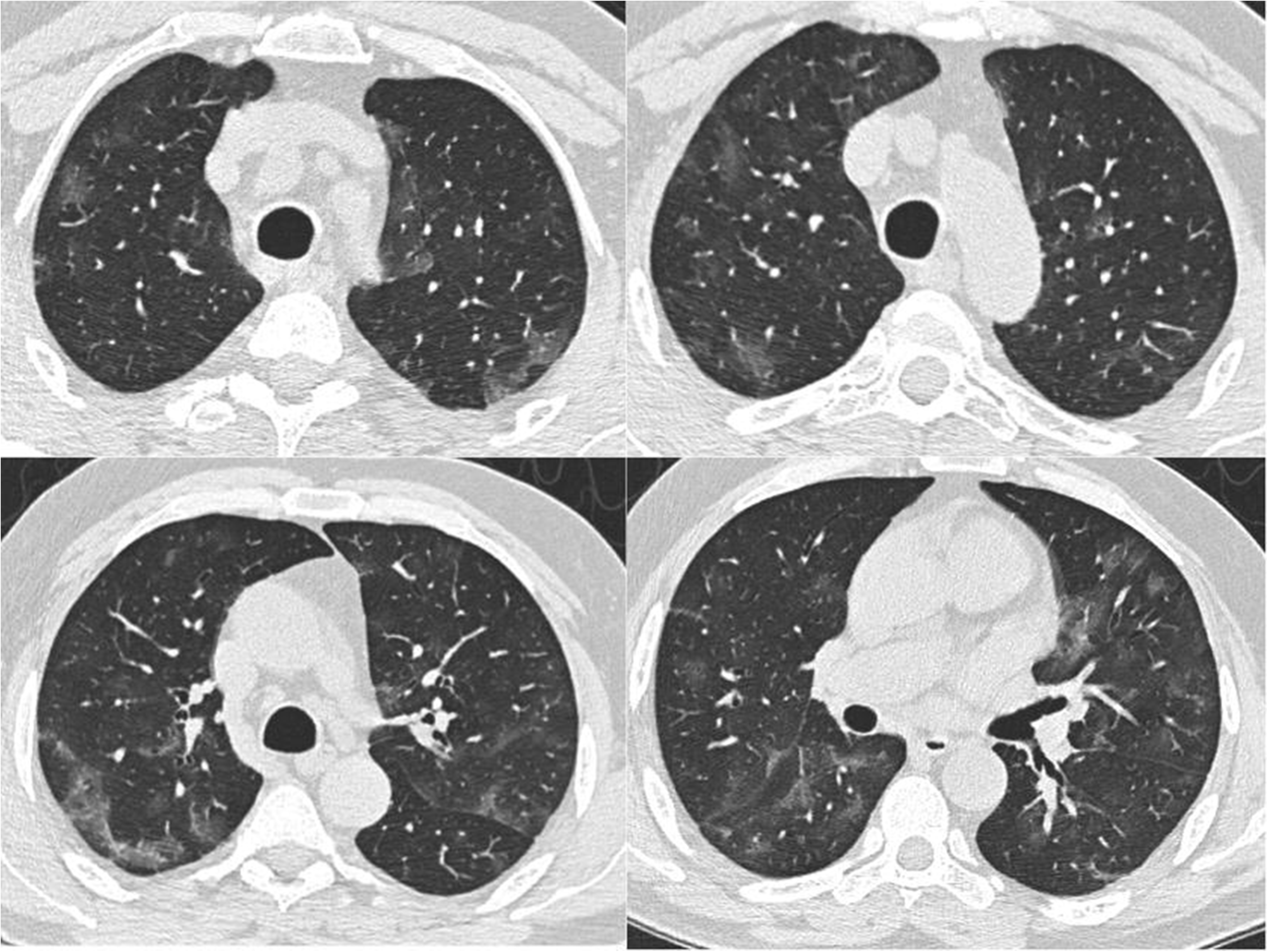
HRCT images showing the bilateral patchy ground-glass opacities.

**Figure 5.**
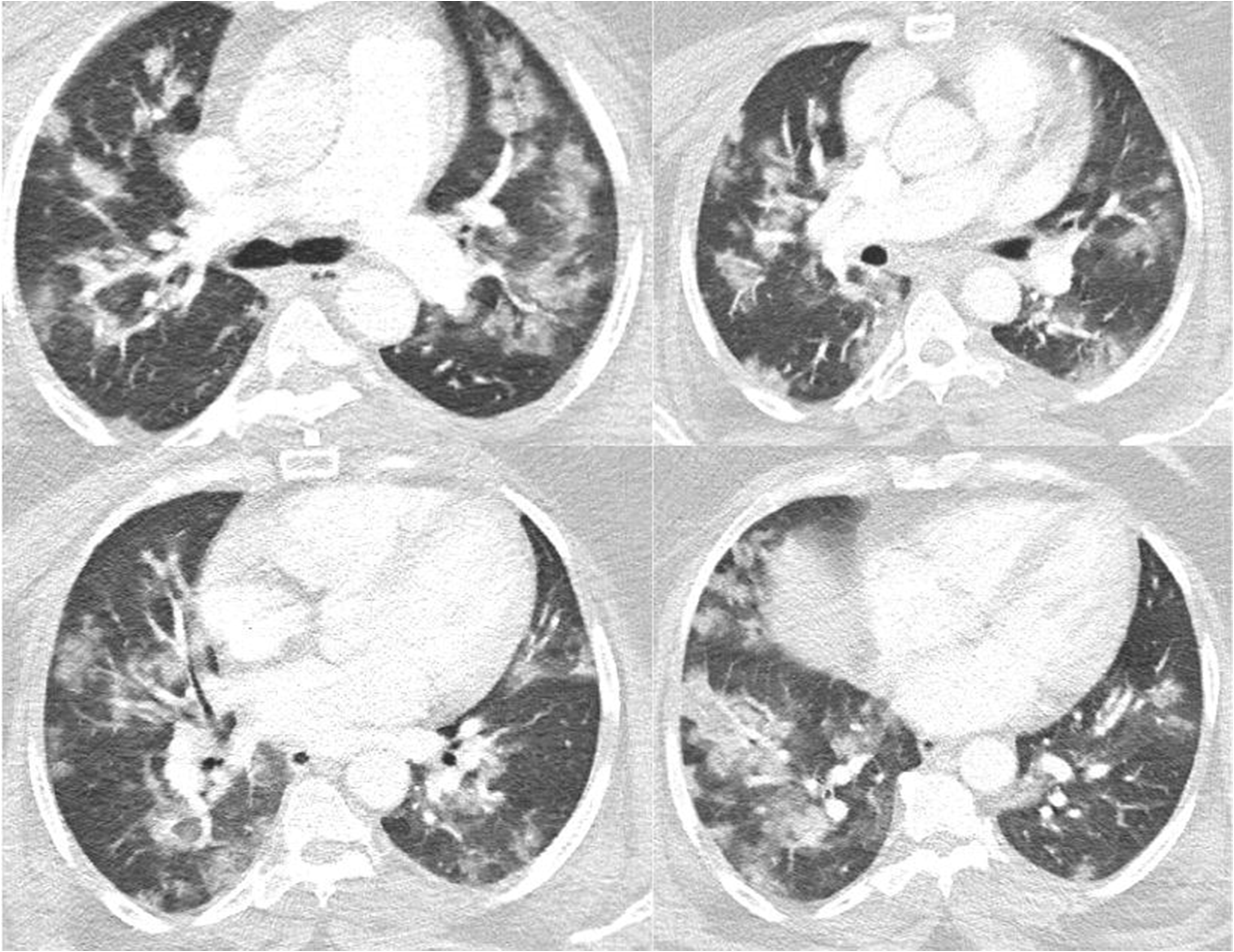
HRCT images showing the bilateral patchy ground-glass opacities with consolidations.

**Figure 6.**
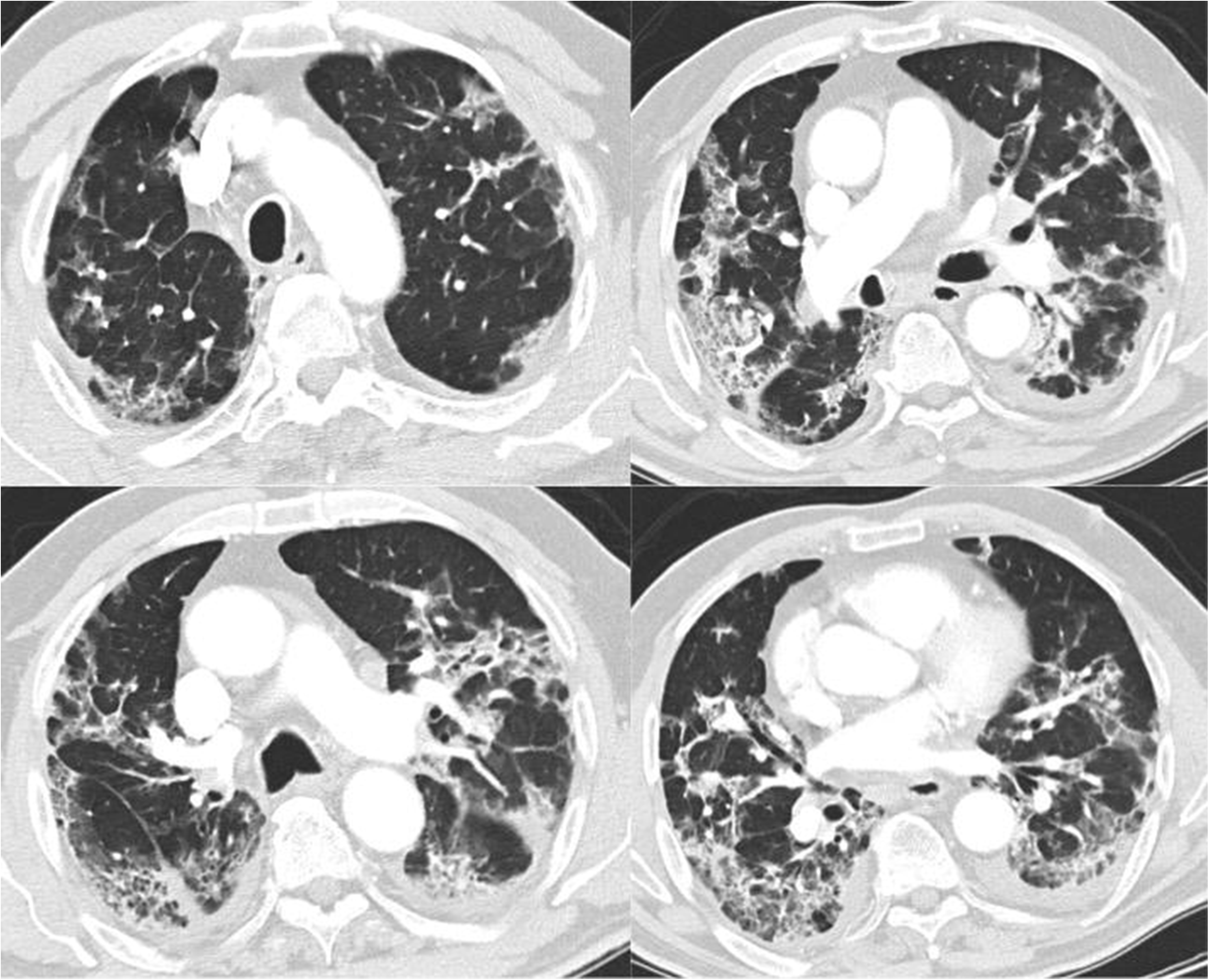
HRCT images showing the bilateral fibroreticular infiltrates with crazy-paving pattern.

## Discussion

In patients with suspected COVID-19 pneumonia, rapid identification, isolation and treatment of infected individuals are crucial. Currently, COVID-19 diagnosis is made using clinical, laboratory and radiological features of patients in the shortest time possible. According to the first COVID-19 pneumonia case series by Bai et al.; the sensitivity of CT is estimated to be 97% compared to PCR tests which have 71% sensitivity^4^. Ai et al. also reported the sensitivity of the RT-PCR test to be in the range of 60% to 70%^5^. Here, our results supported that chest CT results were more sensitive for the detection of COVID-19.

It was suggested that PCR-negative cases with positive CT findings and high clinical suspicion may benefit from repeated PCR testing.^6^ Shi et al. reported that the COVID-19 pneumonia might manifest with chest CT imaging abnormalities, even in asymptomatic patients, with rapid evolution from focal unilateral to diffuse bilateral ground glass opacities that progressed to or coexisted with consolidations within 1–3 weeks. Therefore, combined assessment of imaging features with clinical and laboratory findings could facilitate early diagnosis of COVID-19 pneumonia.^7^ Since radiological evaluation of the thorax is often the key diagnostic element in patients with suspected COVID-19 pneumonia, like in our present study, the patients with typical CT findings but negative RT-PCR results should be isolated and re-evaluated.^8,9^ Furthermore, we think that the diagnosis of COVID-19 should be confirmed by rapid antibody test at least 5 days after the treatment for patients with typical HRCT findings but negative RTPCR results.^9^

SARS-CoV-2 can be detected in different tissues and body fluids. In our study, the samples were taken from the patients with nasopharyngeal and nasal swabs and these samples were run by RT-PCR test. In a study on 1,070 specimens collected from 205 patients with COVID-19, bronchoalveolar lavage fluid specimens showed the highest positive rates (14 of 15; 93%), followed by sputum (72 of 104; 72%), nasal swabs (5 of 8; 63%), fibrobronchoscopy brush biopsy (6 of 13; 46%), pharyngeal swabs (126 of 398; 32%), feces (44 of 153; 29%), and blood (3 of 307; 1%). None of the 72 urine specimens tested positive.^9^ That study showed that sensitivity of nasal and nasopharyngeal swabs for PCR tests remains questionable.

The first larger study on the host humoral response against SARS-CoV-2 has shown that humoral response to SARS-CoV-2 can aid to the diagnosis of COVID-19, including those subclinical cases. In that study, IgA, IgM and IgG response using an ELISA-based assay on the recombinant viral nucleocapsid protein was analyzed in 208 plasma samples from 82 confirmed and 58 probable cases^10,11^. The median duration of IgM and IgA antibody detection were 5 days (IQR 3-6), while IgG was detected on day 14 (IQR 10-18) after symptom onset, with a positive rate of 85.4%, 92.7% and 77.9% respectively. It was shown that detection efficiency by IgM ELISA was higher than that of PCR after 5.5 days of onset of symptoms. In another study of 173 patients, the seroconversion rates (median time) for IgM and IgG were 82.7% (12 days) and 64.7% (14 days), respectively. We noted the mean duration of times from symptoms to hospital admission, hospital stay, drug using and antibody positivity from first symptoms started were 8.6 ± 7.2, 11.2 ± 5.4, 7.9 ± 3.2 and 24 ± 17 days, respectively. It was also reported that a higher titter of antibody was independently associated with severe course of diseases.^12^ Since our study included only PCR-negative COVID-19 patients who were diagnosed and treated based on radiological findings, we were unable to compare the severity of PCR-positive and PCR-negative COVID-19 patients.

A study of 1099 patients in China revealed that 56% of patients had ground-glass opacities, but no radiological findings were reported in 18% of COVID-19 cases. Although bilateral and peripheral ground-glass opacities constitute the most typical CT findings, they are not specific for the COVID-19 disease. Ground glass opacities are also a finding in 33% of paediatric patients. Therefore, we suggest that in PCR-negative cases, radiological diagnosis should be supported with specific antibody detection as seen in our patients.^9,13,14^

Our study has several limitations, including low sample size, the lack of control group and its retrospective nature, however; ideal research conditions are often difficult to be establish during a pandemic situation.

In conclusion, our study remarks the importance of antibody testing in patients which could not be proved with PCR testing for several reasons; such as false negative results or viral clearance of the upper respiratory tract. Even though there is no specific treatment for COVID-19, it is highly important to confirm the diagnosis of highly suspected cases and to prevent further contamination since the retard of diagnosis aided the treatment. Our study demonstrates the feasibility of COVID-19 diagnosis based on rapid-card test for constantly PCR-negative suspected COVID-19 patients. We suggest that detection of antibodies against SARS-CoV-2 with rapid-card test should be included in the diagnostic algorithm in PCR-negative patients with COVID-19 pneumonia.

## Data Availability

The data used in this study is available upon request.

## Acknowledgement

We thank to our colleague Ali Ayata, MD, Assoc. Prof., who provided insight and expertise that greatly assisted the research, although he may not agree with all of the interpretations/conclusions of this paper.

## Author Contributions

Guarantors of integrity of entire study; all authors,

Study concepts/study design or data acquisition or data analysis/interpretation; all authors; manuscript drafting or manuscript revision for important intellectual content; all authors; approval of final version of submitted manuscript; all authors.

## Declaration of Competing Interest

All authors declare that they have no conflict of interest.

## Funding

None.

## Notes

**Declaration of conflict of interest:** None

### Competing Interest Statement

The authors have declared no competing interest.

### Funding Statement

No funding has been received.

### Author Declarations

This is a retrospective study that was specifically approved by the health ministry of Turkey

